# Risk Factors for *Klebsiella* infections among hospitalized patients with Pre-Existing Colonization

**DOI:** 10.1101/2021.02.23.21251995

**Authors:** Krishna Rao, Alieysa Patel, Yuang Sun, Jay Vornhagen, Jonathan Motyka, Abigail Collingwood, Alexandra Teodorescu, Lili Zhao, Keith S. Kaye, Michael Bachman

**Affiliations:** Infectious Diseases Division, Department of Internal Medicine, Michigan Medicine, Ann Arbor, MI; Department of Pathology, Michigan Medicine, Ann Arbor, MI; Department of Biostatistics, School of Public Health, Ann Arbor, MI; Department of Microbiology and Immunology, Michigan Medicine, Ann Arbor, MI

**Author notes:** Corresponding Author: 7510E MSRB 1, 1150 W Medical Center Drive, Ann Arbor, MI 48109.

## Abstract

**Background:** *Klebsiella* commonly colonizes the intestinal tract of hospitalized patients and is a leading cause of healthcare-associated infections. Colonization is associated with subsequent infection, but the factors determining this progression are unclear.

**Methods:** Intensive care and hematology/oncology patients were screened for *Klebsiella* colonization by rectal swab culture and monitored for infection for 90 days after a positive swab. Electronic medical records were analyzed for patient factors associated with subsequent infection, and variables of potential significance in bivariable analysis were used to build a final multivariable model. Concordance between colonizing and infecting isolates was assessed by *wzi* capsular gene sequencing.

**Results:** Among 2087 hospital encounters from 1978 colonized patients, 90 cases of infection (4.3%) were identified. Mean time to infection was 20.6 ±24.69 (range 0-91, median 11.5) days. Of 86 typed cases, 68 unique *wzi* types were identified and 69 cases (80.2%) were colonized with an isolate of the same type prior to infection. Based on multivariable modeling, overall comorbidities, depression and low albumin level at the time of rectal swab were independently associated with subsequent *Klebsiella* infection.

**Conclusions:** Despite the high diversity of colonizing strains of *Klebsiella*, there is high concordance with subsequent infecting isolates and progression to infection is relatively quick. Readily accessible data from the medical record could be used by clinicians to identify colonized patients at increased risk of subsequent *Klebsiella* infection.

**Importance:** *Klebsiella* is a leading cause of healthcare-associated infections. Patients who are intestinally colonized with *Klebsiella* are at significantly increased risk of subsequent infection, but only a subset of colonized patients progress to disease. Colonization offers a potential window of opportunity to intervene and prevent these infections, if the patients at greatest risk could be identified. To identify patient factors associated with infection in colonized patients, we studied 1978 colonized patients. We found that patients with a higher burden of underlying disease in general, depression in particular, and low albumin in a blood test were more likely to be a case of infection. However, these variables did not completely predict infection, suggesting that other host and microbial factors may also be important. The average time to infection was 3 weeks, suggesting that there is time to intervene and prevent *Klebsiella* infections in hospitalized patients.

## Introduction

*Klebsiella pneumoniae* is a leading cause of healthcare-associated infections, commonly causing pneumonia, bacteremia, and urinary tract infections (1). With advancements in genomics, it is clear that infections attributed to *K. pneumoniae* may be caused by related members of a multi-species complex (henceforth *Klebsiella*). Clinical laboratories can now identify *K. variicola,* which may have clinical characteristics distinct from *K. pneumoniae*, but not other complex members such as *K. quasipneumoniae* (2, 3). *Klebsiella* is also a common colonizer of the gastrointestinal tract (4), creating a reservoir of potential disease-causing bacteria. In fact, among colonized patients who develop *Klebsiella* infections, the majority of isolates match those found in the gastrointestinal tract (5, 6). This concordance between colonizing and invasive isolate can be measured by sequencing of the *wzi* capsular gene with the same discriminatory power as 7-gene multi-locus sequence typing (6). This tight link between colonization and infection suggests an opportunity for prevention of infection in hospitalized patients, where detection of colonization could trigger an intervention.

Despite the strong link between colonization and infection, the risk factors for infection in colonized patients are unclear. A combination of patient and bacterial variables likely determines if a patient will develop a *Klebsiella* infection. At a population level without assessment of colonization, increased age, male sex, dialysis, chronic liver disease, solid organ transplant, and cancer were associated with *Klebsiella* bacteremia (7). *Klebsiella* colonization itself is associated with advanced age as well as vancomycin-resistant enterococci (VRE) colonization (8). We previously found certain *Klebsiella* genes and patient factors associated with *Klebsiella* infection compared to asymptomatic colonization (9). However, this study was limited by a small sample size and unknown colonization status in some infected patients.

The goal of this study was to identify risk factors for *Klebsiella* infection among colonized patients at the time of detectable colonization by a rectal swab. In a cohort study of over 2000 hospital encounters with colonized patients, the bacterial species and concordance with subsequent infecting strains was determined. Patient variables were assessed in bivariable analysis and used to create an explanatory model of *Klebsiella* infection. Our findings indicate that readily accessible chart variables can be used to stratify risk of *Klebsiella* infection in colonized patients.

## Results

### Swab collection and *Klebsiella* colonization

From 5/1/2017 to 12/13/2018, rectal swabs collected for VRE screening were cultured for *Klebsiella*, identifying 2098 positive swabs from 1978 patients (recruitment goal=2000). Eliminating duplicate swabs from the same patient within 90 days of the enrollment swab, the cutoff for a subsequent infection to meet our case definition,, resulted in a final cohort consisting of 2087 inpatient admissions with baseline *Klebsiella* colonization among 1978 patients. There was a noticeable seasonal variation in rates of colonization (**Figure 1**) that appeared to co-vary with the mean daily temperature with increased colonization rates occurring in warmer months. Up to 3 isolates were archived from each colonized patient, yielding 5463 *Klebsiella* isolates. Of 2087 patient encounters, 1712 admissions (82.03%) were colonized with *K. pneumoniae* only, 308 (14.76%) were colonized with *K. variicola* only, and 67 (3.21%) admissions were colonized with both *K. pneumoniae* and *K. variicola*.

**Figure 1:**
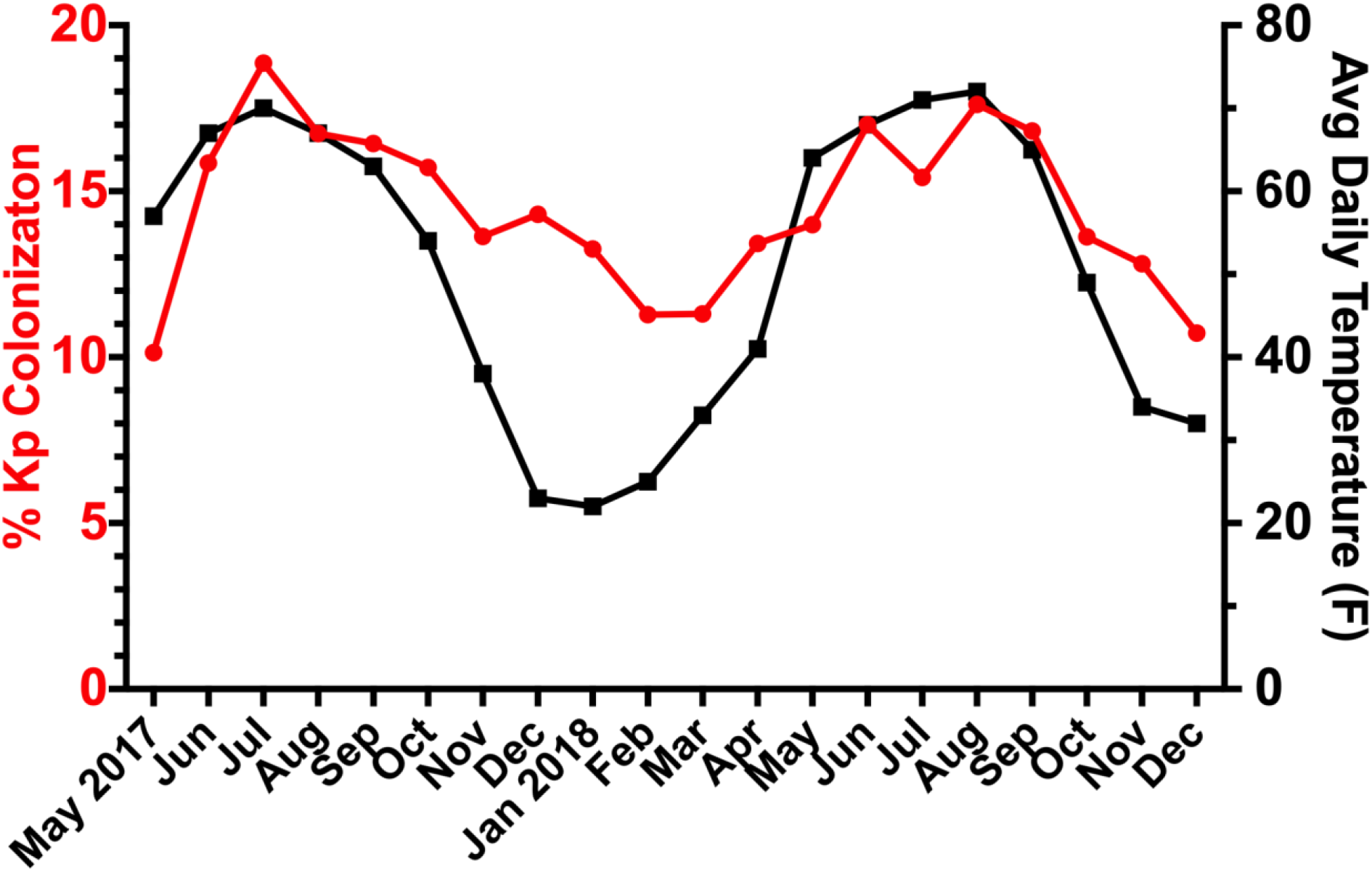
Colonization rate and average daily temperature by month of enrollment. For each month of the study, the percentage of rectal swabs positive for *Klebsiella* is shown, along with the average daily temperature for that month.

### Clinical cultures and case determinations

There were 402 clinical cultures positive for *Klebsiella* from 262 patients, of which 386 (95.8%) were *K. pneumoniae* and 16 (3.97%) were *K. variicola.* These cultures were assessed for cases of *Klebsiella* infection based on chart review. 52 blood cultures were reviewed, and 51 (98.1%) were considered unique cases of infection (one subject had two positive blood cultures and only the first was counted), of which 37 met the 90-day cutoff and were not duplicates (Table 1). 199 urine cultures were reviewed and 46 (23.1%) were considered cases of infection, 22 of which met the 90-day cutoff and were not duplicates. 72 respiratory cultures were reviewed and 40 (55.6%) were considered cases of infection, of which 27 met the 90-day cutoff and were not duplicates. 79 cultures from other body sites were reviewed and 15 (18.8%) were considered cases of infection, of which 4 met the 90-day cutoff and were not duplicates (1 abscess, 2 bile, and 1 tissue). In total, there were 90 infections meeting case definitions for a unique episode within 90 days of the enrollment swab (1 infection occurred 91 days from the index swab but was still included in the analysis), This represents a rate of infection of 4.3% out of 2087 hospital encounters from 1978 *Klebsiella* colonized patients. Bloodstream infections were most frequent (37, 41.1%) followed by respiratory (27, 30.0%), urine (22, 24.4%) and other sites (4, 4.4%).

**Table 1.**
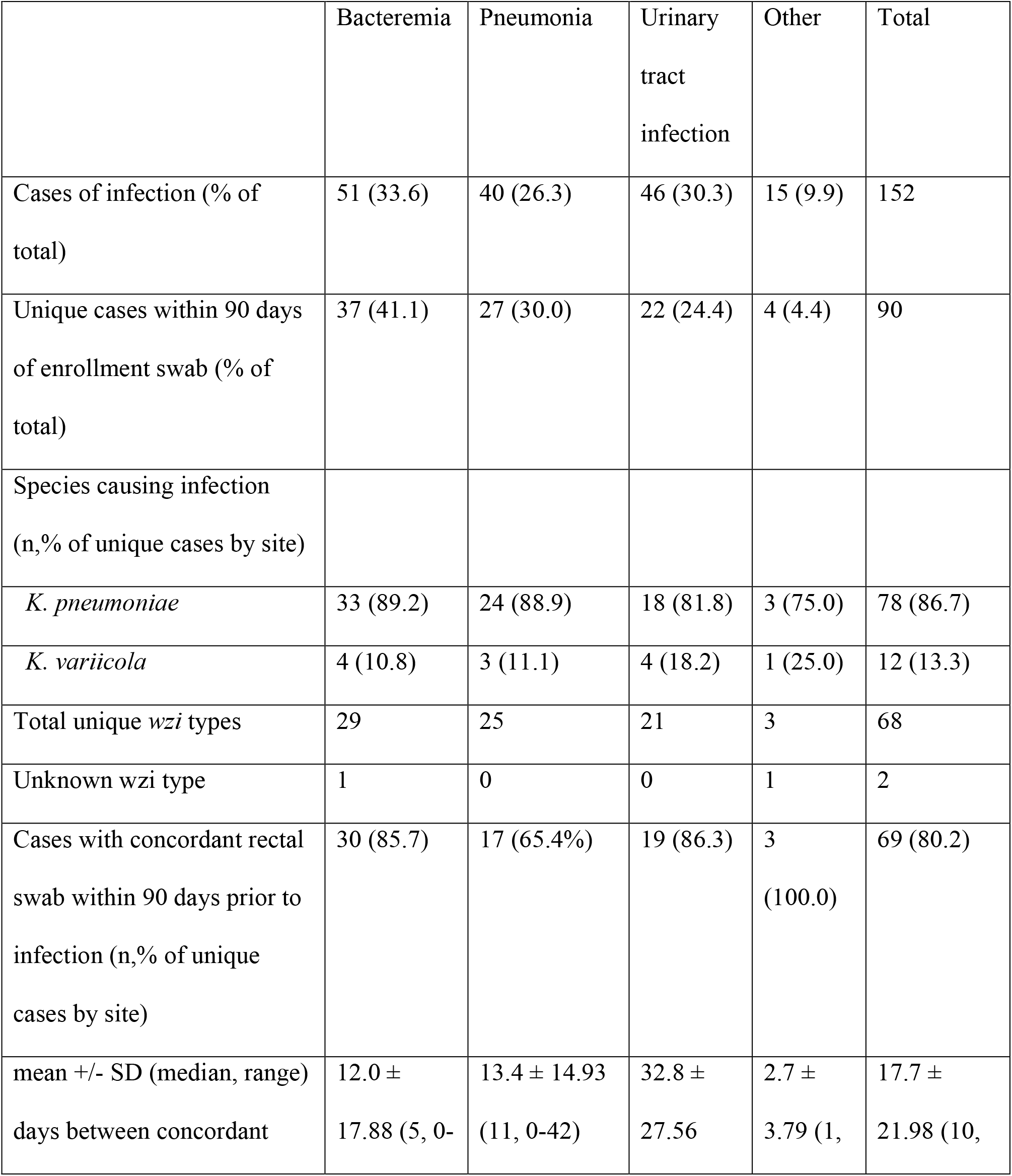
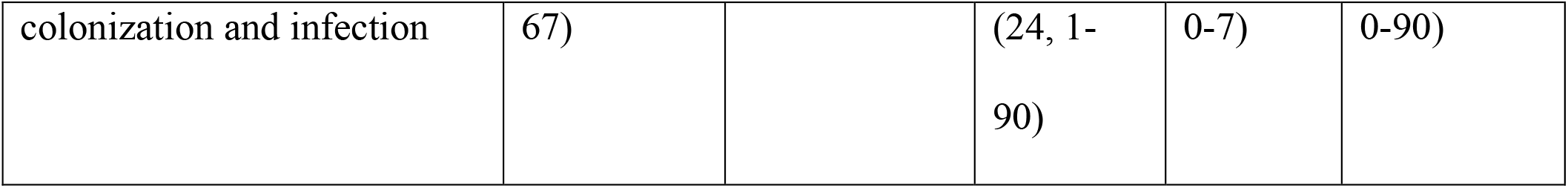
Summary of *Klebsiella* infections by species and strain type.

Most infections were caused by *K. pneumoniae* (86.7%). However, there was high strain diversity with 68 unique *wzi* types observed across 90 infections. Despite this diversity, the majority of these infections were concordant with a *Klebsiella* isolate from the enrollment swab (69/86, 80.2%, 4 cases had missing wzi data). Urinary tract infections (UTI) had the highest rate of concordance (86.3%), bacteremia had a similar rate (81.1%) and pneumonia was lowest (63%; *P*=0.11) The mean time from swab to infection was 20.6 ± 24.69 (median 11.5, range 0-91) for all infections and 17.7 days ± 21.98 (median 10, range 0-90) for concordant cases (**Table 1**). The time to concordant infection (**Figure 2**) differed by site of infection overall (*P* =.001) and was significantly longer for UTI (32.8 ± 27.56) than for bacteremia (12.0 ± 17.88; *P*<0.001) or pneumonia (13.4 ± 14.93; *P*=0.006).

**Figure 2.**
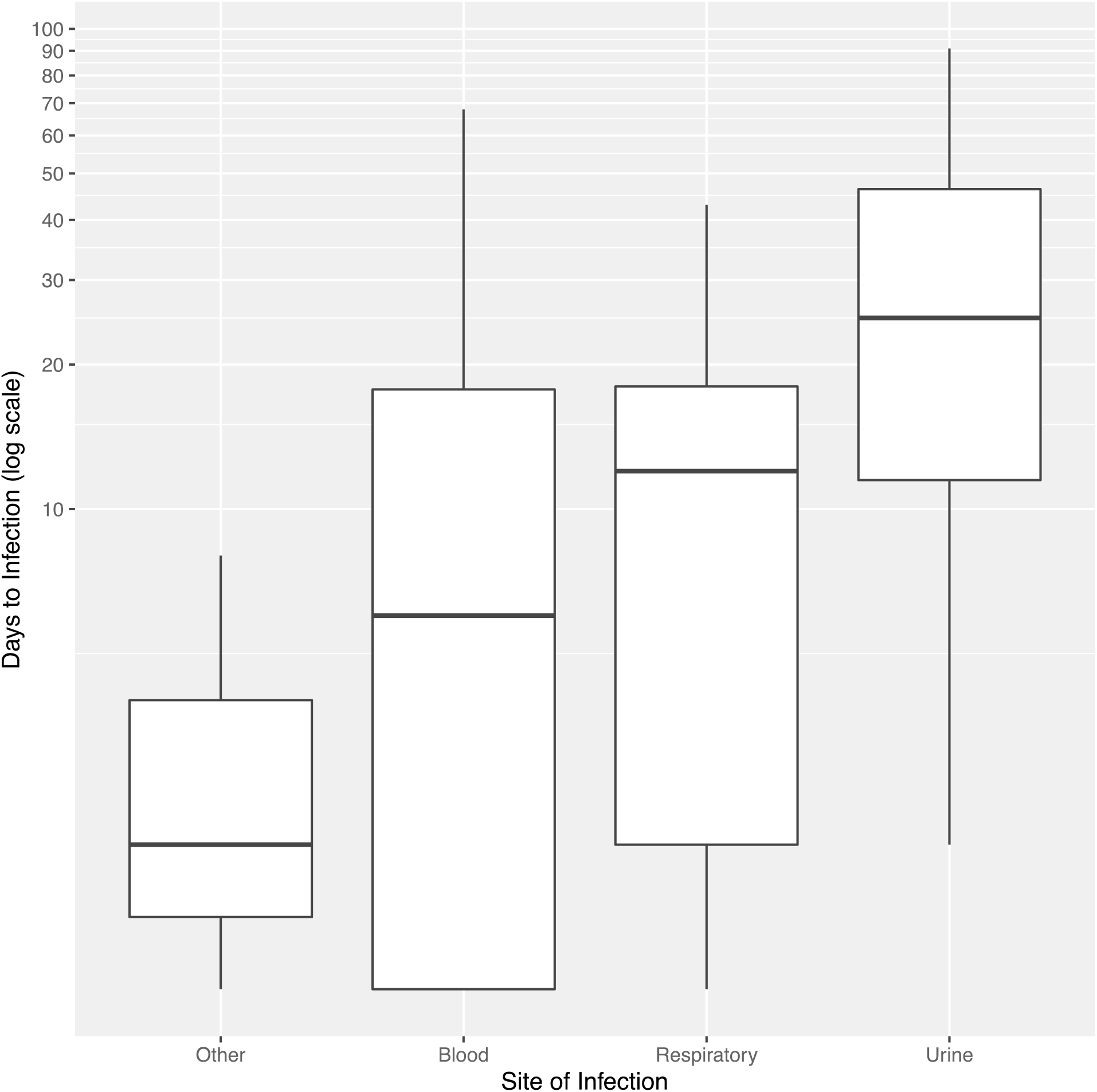
Boxplots of time to concordant infection vs. site of infection. Given the significant right skew of the data and for better visibility, the number of days is plotted on a log scale. The boxes show the medians and interquartile ranges (IQRs), while the whiskers show 1.5x the IQRs.

Among all cases of infection, 19 (21.1%), 13 were found to be extended spectrum beta-lactamase positive (7 bloodstream infections, 4 respiratory, and 2 UTI), and all 13 infecting strains were *K. pneumoniae*. In addition, 6 cases were carbapenem resistant, including 3 bloodstream and 3 respiratory infections. 5 of these strains were *K. pneumoniae* and the other was *K. variicola*. Overall, 17 of these antibiotic-resistant cases were concordant with a colonizing rectal swab.

### Clinical variables and invasive infection with *Klebsiella*

The initial unadjusted analyses compared cases to controls and numerous variables showed statistically significant differences (**Table 2**). There were no significant differences in age, sex, or race between cases and controls. The season that *Klebsiella* colonization was detected on the baseline rectal swab did not associate with increased risk of infection (*P* =0.358). Comorbid illness, as assessed by a Weighted Elixhauser score, and high-risk antibiotic exposure were both enriched in cases compared to controls (**Table 2**). Notably, depression has a neutral weight for predicting in-hospital mortality in the Elixhauser score, but it was significantly associated with *Klebsiella* infection and thus considered separately for the multivariable modeling described below. Invasive devices including urinary catheters and feeding tubes actually had an inverse association with infection and ventilator at baseline trended towards an inverse association (*P*=0.1). A number of medications, including diuretics, antidepressants, Vitamin D, pressors/inotropes, were also associated with *Klebsiella* infection.

**Table 2.**
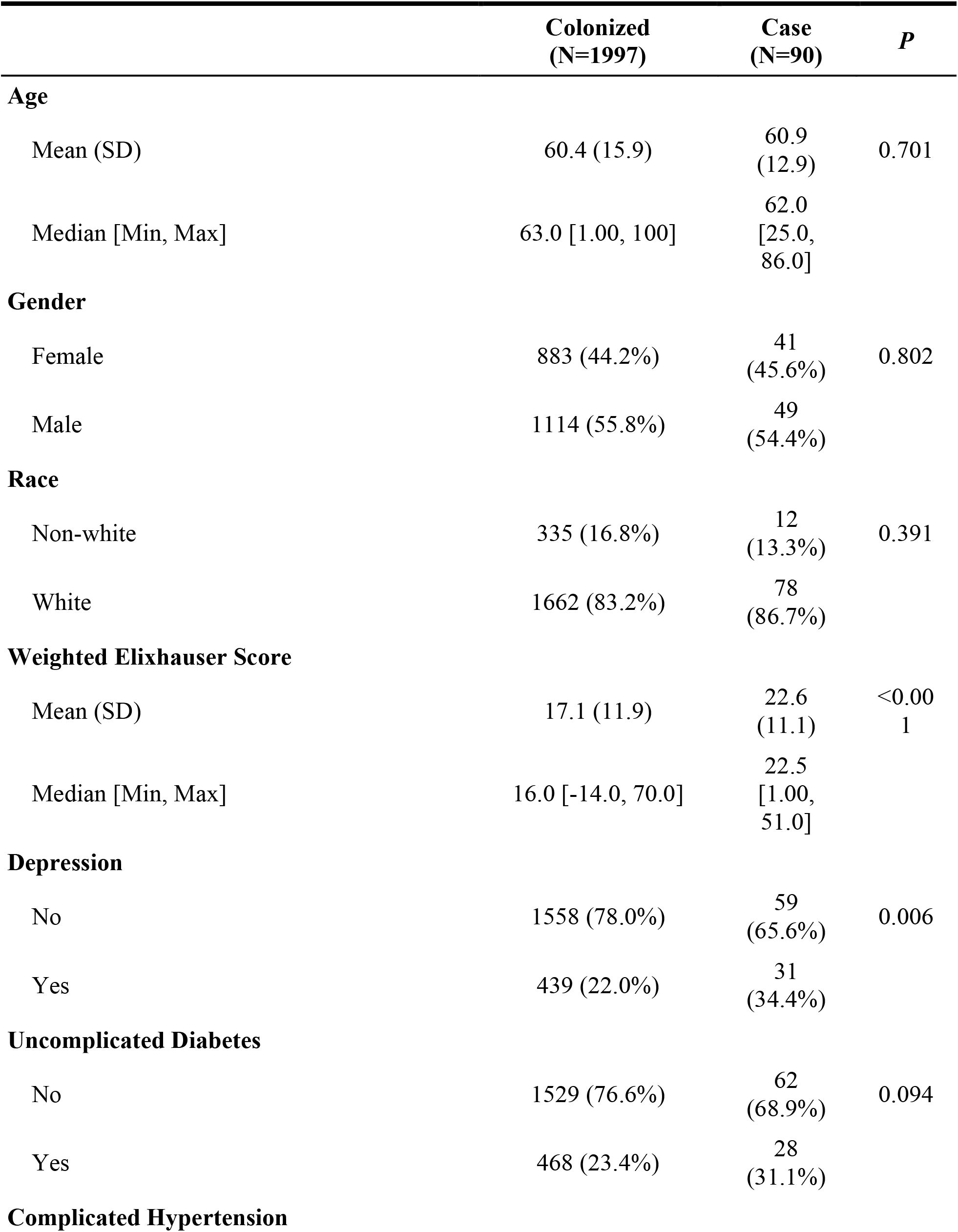
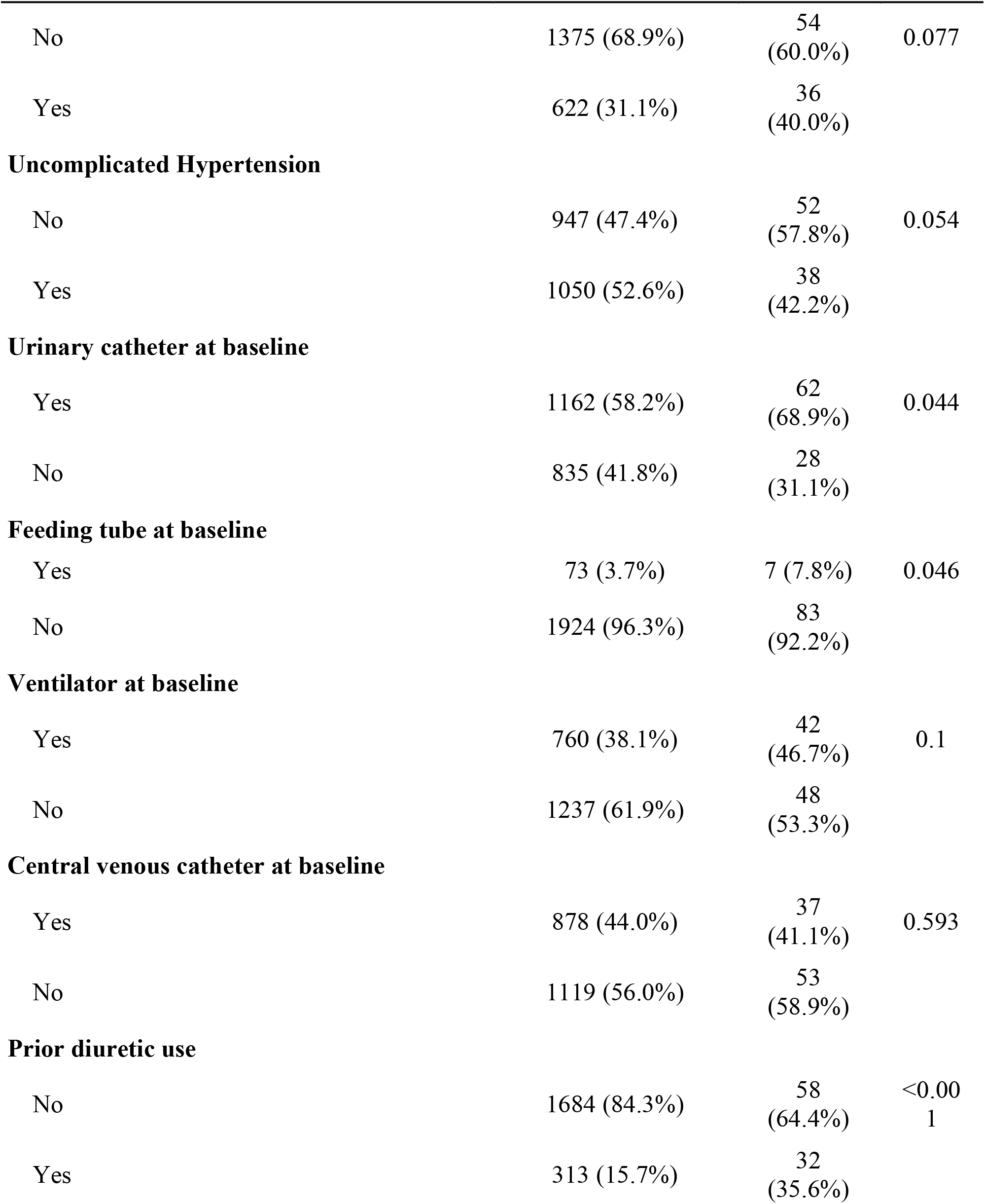
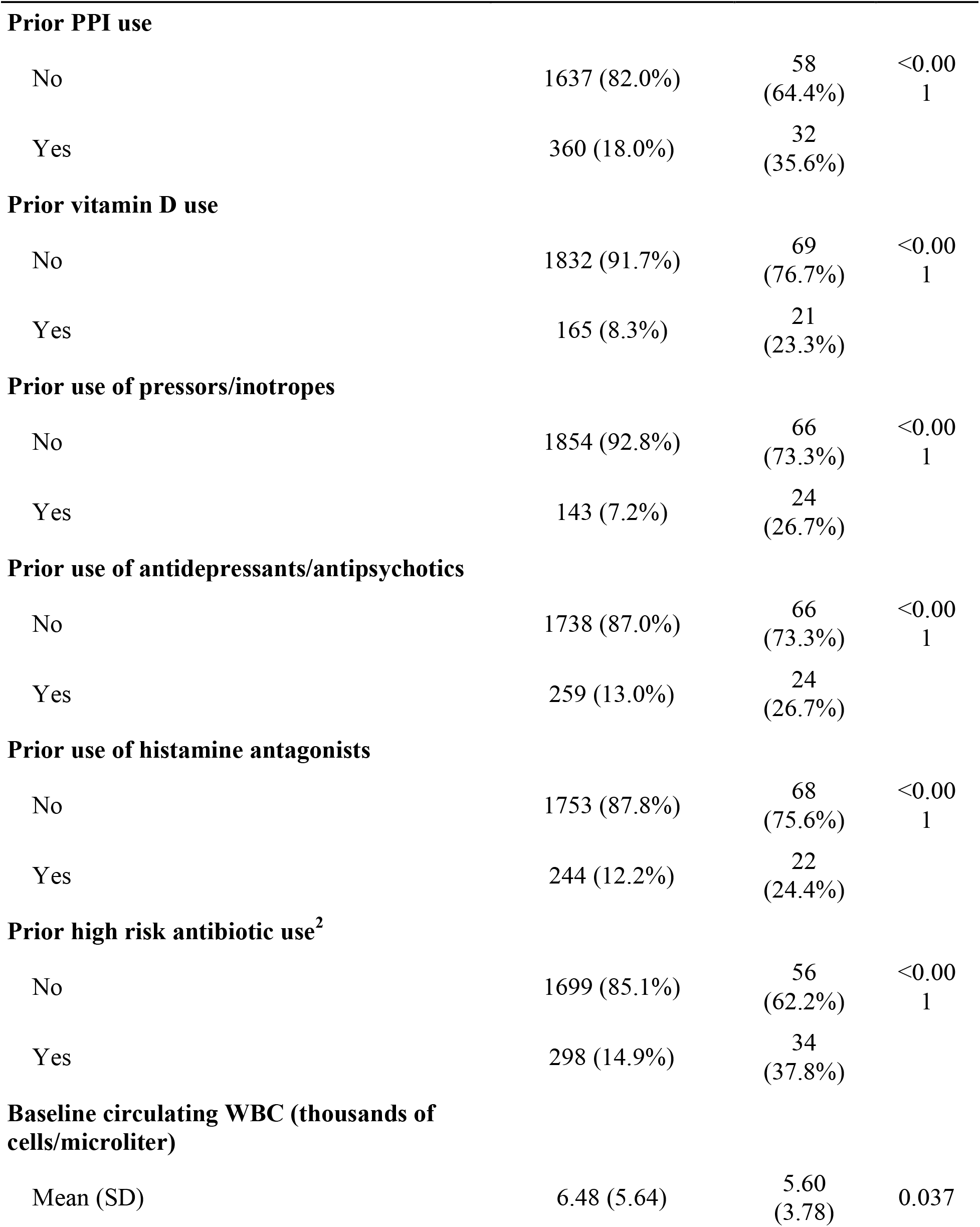
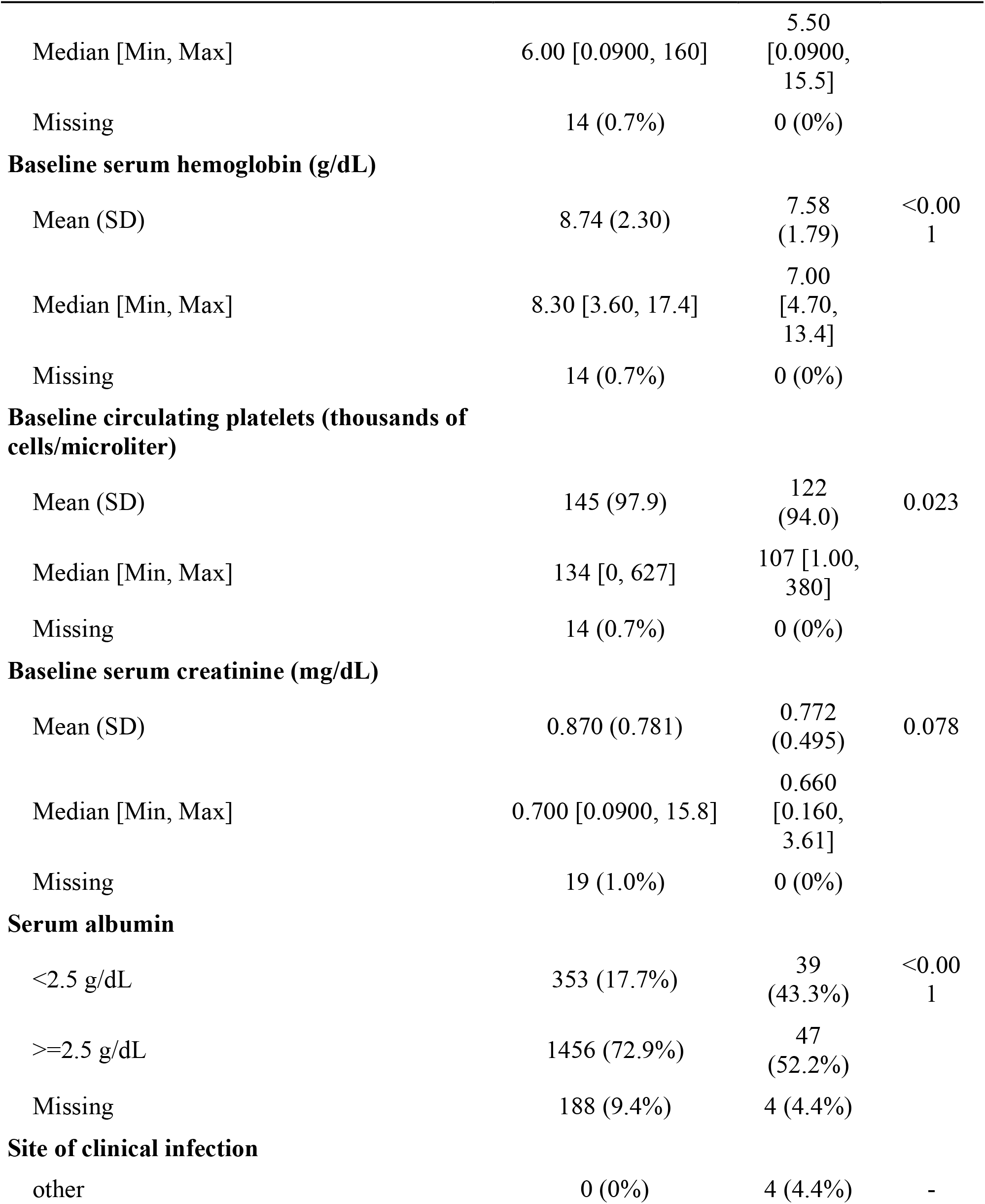
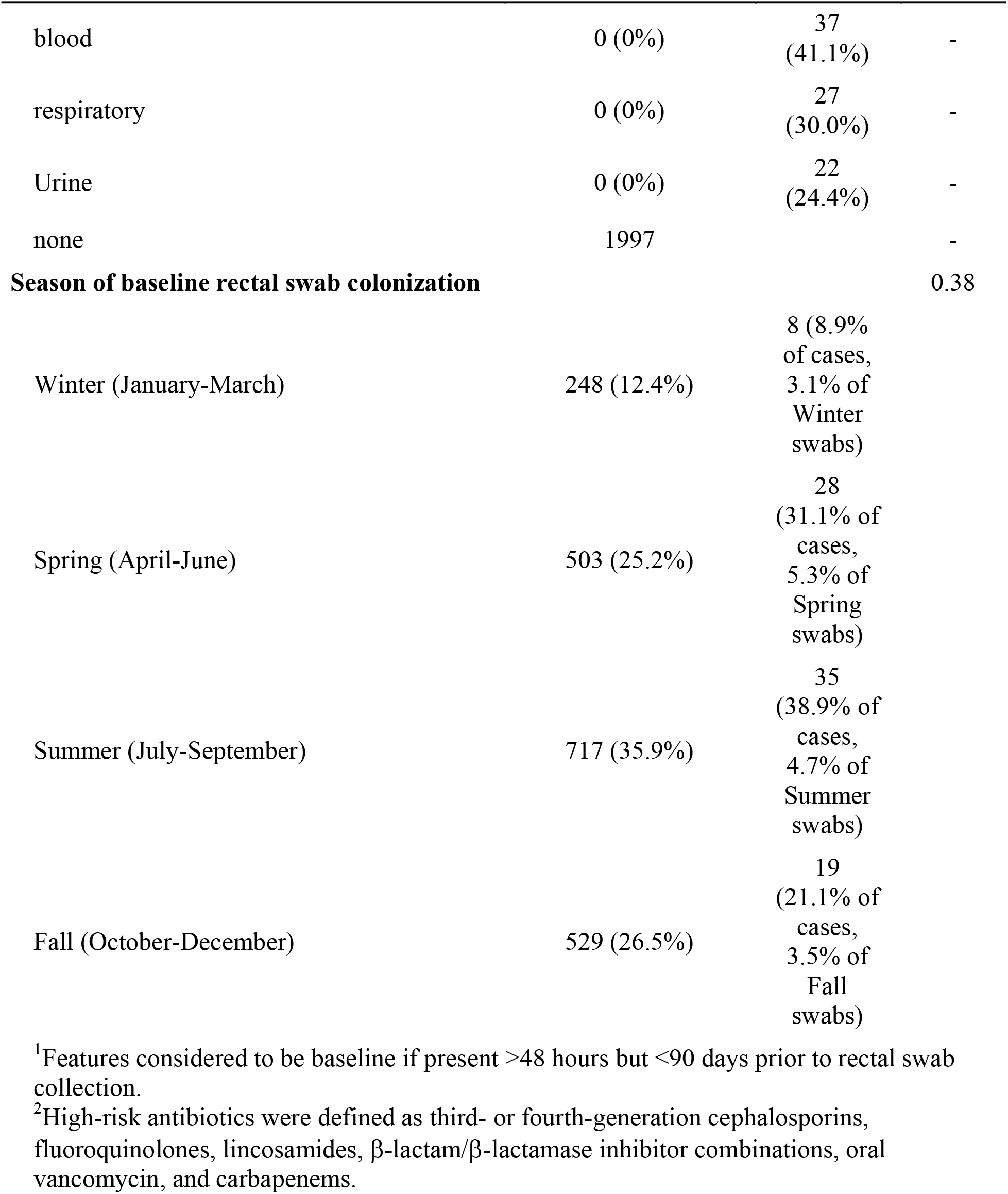
Demographics and selected baseline^1^ characteristics.

### Multivariable Model of *Klebsiella* infection

The purposeful selection process resulted in the explanatory model for *Klebsiella* infection shown in **Table 3**. Here the baseline features of weighted Elixhauser score, depression, and serum albumin <2.5 g/dL associated with an increased risk of subsequent *Klebsiella* infection, and use of pressors/inotropes showed a borderline association with increased risk. Prior diuretic, vitamin D, and high-risk antibiotic use did not independently associate with the outcome, but were retained in the model as confounders. Although depression was selected, anti-depressants were not. The overall fit of the model by area under the receiver operator characteristic curve (AuROC) was 0.74. The mean predicted risk of infection in cases was 8.61% vs. 4.35% in controls, for a discrimination slope of 4.26% (95% confidence interval [CI] 2.59%–5.94%).

**Table 3.** Model of *Klebsiella* infection in colonized patients.

| **Variable** | **OR [95% CI]** | ***P*** |
| --- | --- | --- |
| Elixhauser Score (weighted) | 1.02 [1.00, 1.04] | .034 |
| Depression | 1.80 [1.12, 2.88] | .014 |
| Prior<sup>1</sup> diuretic use | 1.28 [0.70, 2.29] | .407 |
| Prior<sup>1</sup> vitamin D use | 1.36 [0.71, 2.57] | .343 |
| Prior<sup>1</sup> use of pressors/inotropes | 1.9 [0.96, 3.71] | .062 |
| Prior<sup>1</sup> use of high-risk antibiotics | 1.38 [0.74, 2.52] | .298 |
| Albumin < 2.5 g/dL | 2.13 [1.3, 3.48] | .003 |

<sup>1</sup>Features considered to be baseline if present >48 hours but <90 days prior to rectal swab collection.

For the 69 cases of infection that were concordant by *wzi* type with the colonizing isolate, we built a separate multivariable model, shifting the cases without concordance to the control group (**Table 4**). Here the weighted Elixhauser score, depression, and serum albumin <2.5 g/dL were again associated with an increased risk of infection, while baseline serum hemoglobin showed a borderline association such that reduced hemoglobin increased risk. Other potential confounders were not retained. The model AuROC was 0.73, and the mean predicted risk of infection in cases was 5.6% vs. 3.44% in controls for a discrimination slope of 2.16% (95% CI 1.31%–3%). We also conducted a subset analysis by species, and on unadjusted analysis, infection with *K. pneumoniae* vs. *K. variicola* was associated with an increased risk of infection that did not reach statistical significance (OR 1.81 [0.92, 4.1], *P* =.114).

**Table 4.** Model for concordant *Klebsiella* infections (retaining cases without concordance)

| **Variable** | **OR [95% CI]** | ***P*** |
| --- | --- | --- |
| Weighted Elixhauser | 1.03 [1.01, 1.05] | .01 |
| Depression | 2.04 [1.2, 3.41] | .007 |
| Baseline<sup>1</sup> Hemoglobin | 0.88 [0.75, 1.01] | .073 |
| Albumin <2.5 g/dL | 2 [1.15, 3.48] | .013 |

<sup>1</sup>Features considered to be baseline if present >48 hours but <90 days prior to rectal swab collection.

## Discussion

This study of over 2000 hospital encounters provides a comprehensive analysis of the dynamics of *Klebsiella* infection in colonized patients and identifies patient factors that are present at admission and associated with infection. Overall, a small but significant percentage of colonized patients (4.3%) progress to infection. These infections are caused by diverse strains of *Klebsiella* and are predominantly caused by strains present in a rectal swab (80.2%) at the time of admission. When infection occurs in colonized patients, it is within days to weeks (median 11.5 days) from the first positive rectal swab. The risk of infection is influenced, at least in part, by characteristics of the colonized patients themselves. In particular, the overall burden of comorbidities, pre-existing diagnosis of depression, and a low albumin at baseline are significant and independently associated with infection. Identification of colonized patients at high-risk of *Klebsiella* infection at the time of admission could be used to direct interventions to prevent these infections in our most vulnerable patients.

The dynamics of colonization confirm and expand on previous studies showing a strong link between *Klebsiella* colonization and infection. Indeed, in a smaller pilot study we demonstrated that intensive care patients were frequently colonized (23%), colonization was associated with infection (5.2% vs 1.3% uncolonized patients; OR 4.01), the colonizing isolates were diverse, and infections were caused often by colonizing strains (13/16 paired isolates matched) (6). Similarly, Gorrie et al found colonization rates of 5.9-19% among intensive care patients, a strong association between colonization and infection (OR 6.9; 13 infections among 27 colonized patients) and high diversity of colonizing and infecting isolates (5). The current study focusing on colonized patients refines the results of these smaller studies, demonstrating a similar rate of infections (4.3%), high diversity of infecting strains and a more accurate estimate of concordant infections (80.2%). It also demonstrates that detectable colonization does not capture all of the infection risk, and that patient variables modify this risk.

The predominance of concordant infections among colonized patients, and the fact that the infecting strain is often present at admission, indicates that infection prevention for *Klebsiella* should focus on endogenous, patient risk factors. To that end, our study determined that comorbidities, medication history, and routine laboratory results at admission are informative regarding infection risk, with cases having twice the predicted risk compared to controls (8.61% vs. 4.35%). Importantly, the model for concordant infection (**Table 4**) was more parsimonious than the model for all infections (**Table 3**). Infection risk increased with the Elixhauser comorbidity score, and depression contributed substantially (OR 2.04, CI 1.2-3.41; *P* =0.007). While it is not clear in our study if the association is causal, exposure to antidepressant medications is known to significantly alter the gut microbiome (10). Additionally, prior research has identified depression and antidepressant medication use as risk factors *C. difficile* infection, an infection that is mediated through disruption of the gut microbiome (11). Although antidepressant use was associated with *Klebsiella* infection on unadjusted analysis, it was confounded by other variables. When we added antidepressant use to the final model, it was not statistically significant (P=.66), nor was it when we replaced depression with antidepressant use in the model (P=0.3). It is plausible that the gut microbiome mediates the association between depression and subsequent *Klebsiella* infection that we observed in colonized patients, but this requires further study.

The association between low albumin and infection (OR 2.0, CI 1.15-3.48, P=0.013) was also previously observed for *Klebsiella* infections in the ICU including both colonized and non-colonized patients (6). Low albumin can be caused by decreased synthesis due to malabsorption, malnutrition or hepatic dysfunction, or due to losses from ascites, nephropathy, or enteropathy (12). Although the pathophysiologic link between low albumin and *Klebsiella* infection is unclear, this is an informative and readily widely available predictor, ordered by the physician in > 90% of the cohort.

In this multi-year study, the rates of colonization wax and wane by month with the highest rates in the summer. This seasonal pattern was previously demonstrated for *Klebsiella* bacteremia across 4 continents (13), with both higher temperature and humidity being associated with increased rates. The observed seasonality of *Klebsiella* colonization may in turn cause a seasonality of infection. Indeed, the proportion of cases by season varied from a low of 8.9% in Winter to a high of 38.9% in Summer (**Table 2**), but tracked closely with the overall proportion of colonized patients by season and the risk of infection in a given colonized patient did not vary by season (*P* =0.358). The seasonality of *Klebsiella* bacteremia is not seen for the related pathogens *Escherichia coli* or *Serratia* (13). Therefore, prevention efforts that focus on identifying environmental reservoirs of *Klebsiella* that fluctuate with temperature could have a significant impact on overall rates of infection through reduction of the initial colonization event rate.

Our study has several limitations including its retrospective design focused on a convenience sample of ICU/oncology ward patients with post-hoc data extraction. We undertook several strategies to mitigate this central limitation. We only included variables that we felt sure were baseline features present at colonization and not a reflection of emerging or already present infection (**Supplementary Methods**), and for variables such as vitals, laboratory results, and medication/device exposures we either excluded them from modeling, captured them only if present prior to swab collection, and/or manually confirmed their baseline status by chart review. We also used matching and multivariable modeling to reduce bias/confounding, and we conducted several sensitivity analyses to confirm our main results. We also could not confirm associations for variables that in prior research were found to be associated with *Klebsiella* infection. A population-based study of *Klebsiella* bacteremia found male sex, advanced age, dialysis, solid-organ transplantation, chronic liver disease, and cancer were associated with infection (7). We previously found that advanced age was associated with colonization (8). Like with temperature and seasonality, some infection risks may actually represent colonization risks. Although the comorbidities were not categorized in the same way across studies, none of the other risk factors were significantly associated with infection in colonized patients (**Supplementary Results**). Finally, technical limitations arising from the sensitivity of our plating method and picking 3 colonies per plate for characterization could have led us to miss some patients who were colonized at baseline and to miss a low abundance, concordant strain in infected patients with detectable colonization.

Although the models of *Klebsiella* infection had significant explanatory power, there are likely to be additional factors that modulate the risk of infection in colonized patients. Relative abundance of *Klebsiella* colonization is also associated with infection, even after controlling for the variables identified here (co-submitted manuscript), and it could further refine infection risk in colonized patients. In addition, the complex genomics of *Klebsiella* is likely to be an important predictor of infection, and we have previously identified several putative pathogenicity loci (9). The present study can further inform such future work by identifying the most important clinical variables that should be considered as confounders when assessing the independent contribution of bacterial genetic loci, colonization density and other features of the gut microbial community on risk of subsequent *Klebsiella* infection. Such a holistic approach to modeling the risk of infection, from host characteristics to the gut community to the genetics of the colonizing strain, may be what is needed to develop the most accurate and useful models of invasive *Klebsiella* infection.

## Methods

### Cohort identification, sample collection, and strain typing

This study was approved by the Institutional Review Boards of the University of Michigan Medical School (IRBMED). We sequentially enrolled subjects to establish a cohort of patients with *Klebsiella* (*K. pneumoniae* or *K. variicola*) rectal colonization. Inpatients admitted to the intensive care units [ICUs] and the oncology wards of the University of Michigan hospitals in Ann Arbor, Michigan, from May 2017 to September 2018, were eligible for inclusion if discarded vancomycin resistant *Enterococcus* [VRE] screening rectal swab media was available for culture. All patients admitted to these wards undergo screening for rectal VRE carriage per hospital policy, using a flocked swab placed into Amies transport media (E-swab, Becton Dickinson, Franklin Lakes, NJ). Subjects were subsequently enrolled into our study if *K. pneumoniae* was isolated from the swab media through plating on MacConkey agar followed by taxonomic identification using MALDI TOF mass spectrometry. Up to 3 isolates per rectal swab culture were archived along with all clinical culture isolates positive for *K. pneumoniae*. Wzi typing was done using PCR and Sanger sequencing (14). Wzi type was assigned by uploading the consensus gene sequence to the BigsDB database (https://bigsdb.pasteur.fr).

### Colonization rates by month

The percentage of rectal swabs containing *Klebsiella* was calculated for each month from records of MacConkey agar growth and MALDI-TOF results for species identification. Average daily temperature for the month was obtained for Ann Arbor from www.wunderground.com.

### Clinical data extraction and case definitions

Structured queries were used to abstract clinical data from the electronic medical record including demographics, pre-existing comorbidities, vitals, laboratory results, imaging and procedure reports. These clinical data were used to construct variables comprised of baseline features present at or before the time of swab collection. As described further in our results, certain variables were collapsed into colligative variables for the unadjusted and adjusted analyses. Positive clinical cultures for *Klebsiella* species from the day of, to 90 days after, rectal swab collection were independently reviewed for alignment with our case definitions by two clinician study team members (J.B. and M.B), and disagreements were adjudicated by a third clinician team member (K.R.). All patients with a blood culture growing *Klebsiella* were considered to have an infection. Manual chart review was conducted by the study team to decide if potential cases met published infection criteria from professional societies / governmental organizations for pneumonia or urinary tract infection (15-19), and cultures from other sites, such as wounds, were considered cases based on the clinical judgment of reviewers. For those meeting clinical case definitions of infection, the clinical isolate and preceding rectal swab isolates were evaluated for concordance of *Klebsiella* isolates by *wzi* gene sequencing as previously described (6, 14). We have previously demonstrated that *wzi* sequencing has similar discriminatory power to 7-gene multi-locus sequence typing (6).

### Statistical Methods

Initial descriptive statistics, visualizations such as histograms, and bivariable relationships between predictors and the primary outcome of invasive infection were calculated and used to inform variable construction and inclusion for multivariable modeling. Bivariable relationships were assessed with Student’s t-tests (or ANOVA first if multiple categories) and Chi-squared tests. Given the primary outcome occurring at low frequency and the resulting statistical power considerations, we sought to collapse variables into colligative metrics where applicable. For example, a weighted Elixhauser score (20) was included for modeling in lieu of most individual comorbidities, save when the weight was set to zero or if the weight differed notably in the direction/magnitude from what was observed on bivariable associations with the primary outcome. High-risk antibiotics were defined as third-or fourth-generation cephalosporins, fluoroquinolones, lincosamides, β-lactam/β-lactamase inhibitor combinations, oral vancomycin, and carbapenems, based on their association with disruption of the intestinal microbiome (21). We first constructed an explanatory model to identify independent predictors of the primary outcome. We did this using a modified purposeful selection approach (22) to allow for inclusion of variables in the final model that adjust for confounding in other covariates, even if not independently associated with the primary outcome themselves (see **Supplementary Methods**). Following this, we constructed additional models to test hypotheses about specific risk factors that were not selected by the above procedure and to conduct sensitivity analyses using the primary model from the above procedure. The sensitivity analyses included limiting the case definition to only apply when isolates from clinical infections were concordant with the colonizing strain by *wzi* sequencing; and assessing how colonizing species influences risk of infection. All analyses were conducted in R version 4.0.3 (R Foundation for Statistical Computing, Vienna, Austria). A cutoff *P*-value of <.05 was used for statistical inference. Model performance was further assessed using the area under the receiver operator characteristic curve (AuROC) and the discrimination slope using the R package *pROC* (23).

## Supporting information

Supplemental Methods and Results

STROBE Checklist

## Data Availability

Deidentified data from human subjects may be made available upon request, pending approval from the University of Michigan Institutional Review Board.

## Acknowledgements

Research reported in this publication was supported by the National Institute Of Allergy And Infectious Diseases of the National Institutes of Health under Award Number R01AI125307. The content is solely the responsibility of the authors and does not necessarily represent the official views of the National Institutes of Health.

