## Supplemental Methods and Results for "Risk Factors for *Klebsiella* infections among hospitalized patients with Pre-Existing Colonization"

### Supplementary Materials

### Supplementary Methods

Variable Selection and Modeling. We only considered variables for inclusion if they were significant on the unadjusted analysis and if we were sure they represented baseline features and not the consequences of infection we are trying to model. For example, serum creatinine, circulating platelets, and circulating WBC were not considered for modeling even if significant on unadjusted analysis, since these variables can rise during infection. Additionally, we only counted exposures to devices and medications if we could confirm they were present at baseline (>48 hours but <90 days before the swab collection) and not after the swab was already collected.

The final explanatory model presented was constructed using a purposeful selection approach. Purposeful selection begins with an unadjusted analysis of each variable to select candidates with statistically significant associations with the outcome, and these are included in the starting set of covariates for the multivariable model. Iteratively, covariates are then removed from the model if they are non-significant ($P > .05$) and not a confounder (i.e. do not affect the estimate of other variables' coefficients by at least 20%). A change in a parameter estimate above the specified level indicates that the excluded variable was important in the sense of providing a needed adjustment for one or more of the variables remaining in the model (i.e. it should be retained even if not significant). The resulting model contains significant covariates and other confounders, and then variables not included are added back one at a time. Once again, the model is iteratively reduced as before but only for the variables that were additionally added. At the end of this final step, we are left with a multivariable model for *Klebsiella* infection.

**Supplementary Results**

**Full Unadjusted Result Tables**

Comorbidities and Devices

| | **Colonized (N=1997)** | **Case (N=90)** | ***P*** |
|---|---|---|---|
| **Age** | | | |
| Mean (SD) | 60.4 (15.9) | 60.9 (12.9) | 0.701 |
| Median [Min, Max] | 63.0 [1.00, 100] | 62.0 [25.0, 86.0] | |
| **Gender** | | | |
| Female | 883 (44.2%) | 41 (45.6%) | 0.802 |
| Male | 1114 (55.8%) | 49 (54.4%) | |
| **Race** | | | |
| Non-white | 335 (16.8%) | 12 (13.3%) | 0.391 |
| White | 1662 (83.2%) | 78 (86.7%) | |
| **Weighted Elixhauser Score** | | | |
| Mean (SD) | 17.1 (11.9) | 22.6 (11.1) | <0.001 |
| Median [Min, Max] | 16.0 [-14.0, 70.0] | 22.5 [1.00, 51.0] | |
| **Alcohol Abuse** | | | |
| No | 1886 (94.4%) | 84 (93.3%) | 0.655 |
| Yes | 111 (5.6%) | 6 (6.7%) | |
| **Blood Loss Anemia** | | | |
| No | 1854 (92.8%) | 71 (78.9%) | <0.001 |
| Yes | 143 (7.2%) | 19 (21.1%) | |
| **Cardiac Arrhythmias** | | | |
| No | 979 (49.0%) | 36 (40.0%) | 0.094 |
| Yes | 1018 (51.0%) | 54 (60.0%) | |
| **Chronic Pulmonary Disease** | | | |
| No | 1436 (71.9%) | 56 (62.2%) | 0.047 |
| Yes | 561 (28.1%) | 34 (37.8%) | |
| **Coagulopathy** | | | |
| No | 1432 (71.7%) | 53 (58.9%) | 0.009 |

| | Colonized (N=1997) | Case (N=90) | *P* |
|---|---|---|---|
| Yes | 565 (28.3%) | 37 (41.1%) | |
| **Congestive Heart Failure** | | | |
| No | 1425 (71.4%) | 60 (66.7%) | 0.337 |
| Yes | 572 (28.6%) | 30 (33.3%) | |
| **Iron Deficiency Anemia** | | | |
| No | 1790 (89.6%) | 77 (85.6%) | 0.218 |
| Yes | 207 (10.4%) | 13 (14.4%) | |
| **Depression** | | | |
| No | 1558 (78.0%) | 59 (65.6%) | 0.006 |
| Yes | 439 (22.0%) | 31 (34.4%) | |
| **Uncomplicated Diabetes** | | | |
| No | 1529 (76.6%) | 62 (68.9%) | 0.094 |
| Yes | 468 (23.4%) | 28 (31.1%) | |
| **Complicated Diabetes** | | | |
| No | 1677 (84.0%) | 74 (82.2%) | 0.658 |
| Yes | 320 (16.0%) | 16 (17.8%) | |
| **Drug Abuse** | | | |
| No | 1891 (94.7%) | 86 (95.6%) | 0.72 |
| Yes | 106 (5.3%) | 4 (4.4%) | |
| **Fluid & Electrolyte Disorders** | | | |
| No | 911 (45.6%) | 29 (32.2%) | 0.013 |
| Yes | 1086 (54.4%) | 61 (67.8%) | |
| **Complicated Hypertension** | | | |
| No | 1375 (68.9%) | 54 (60.0%) | 0.077 |
| Yes | 622 (31.1%) | 36 (40.0%) | |
| **Uncomplicated Hypertension** | | | |
| No | 947 (47.4%) | 52 (57.8%) | 0.054 |
| Yes | 1050 (52.6%) | 38 (42.2%) | |
| **Hypothyroidism** | | | |
| No | 1672 (83.7%) | 76 (84.4%) | 0.856 |

| | Colonized (N=1997) | Case (N=90) | *P* |
|---|---|---|---|
| Yes | 325 (16.3%) | 14 (15.6%) | |
| **Liver Disease** | | | |
| No | 1661 (83.2%) | 69 (76.7%) | 0.109 |
| Yes | 336 (16.8%) | 21 (23.3%) | |
| **Lymphoma** | | | |
| No | 1789 (89.6%) | 82 (91.1%) | 0.642 |
| Yes | 208 (10.4%) | 8 (8.9%) | |
| **Metastatic Cancer** | | | |
| No | 1672 (83.7%) | 71 (78.9%) | 0.226 |
| Yes | 325 (16.3%) | 19 (21.1%) | |
| **Obesity** | | | |
| No | 1360 (68.1%) | 59 (65.6%) | 0.612 |
| Yes | 637 (31.9%) | 31 (34.4%) | |
| **Other Neurological Disorders** | | | |
| No | 1696 (84.9%) | 63 (70.0%) | <0.001 |
| Yes | 301 (15.1%) | 27 (30.0%) | |
| **Paralysis** | | | |
| No | 1895 (94.9%) | 84 (93.3%) | 0.514 |
| Yes | 102 (5.1%) | 6 (6.7%) | |
| **Peptic Ulcer Disease Excluding Bleeding** | | | |
| No | 1939 (97.1%) | 84 (93.3%) | 0.043 |
| Yes | 58 (2.9%) | 6 (6.7%) | |
| **Peripheral Vascular Disorders** | | | |
| No | 1603 (80.3%) | 74 (82.2%) | 0.648 |
| Yes | 394 (19.7%) | 16 (17.8%) | |
| **Psychoses** | | | |
| No | 1943 (97.3%) | 85 (94.4%) | 0.11 |
| Yes | 54 (2.7%) | 5 (5.6%) | |
| **Pulmonary Circulation Disorders** | | | |
| No | 1641 (82.2%) | 73 (81.1%) | 0.797 |

| | Colonized (N=1997) | Case (N=90) | *P* |
|---|---|---|---|
| Yes | 356 (17.8%) | 17 (18.9%) | |
| **Renal Failure** | | | |
| No | 1483 (74.3%) | 61 (67.8%) | 0.17 |
| Yes | 514 (25.7%) | 29 (32.2%) | |
| **Rheumatoid Arthritis & Collagen Vascular Diseases** | | | |
| No | 1878 (94.0%) | 82 (91.1%) | 0.255 |
| Yes | 119 (6.0%) | 8 (8.9%) | |
| **Solid Tumor Without Metastasis** | | | |
| No | 1521 (76.2%) | 62 (68.9%) | 0.115 |
| Yes | 476 (23.8%) | 28 (31.1%) | |
| **Valvular Disease** | | | |
| No | 1590 (79.6%) | 82 (91.1%) | 0.008 |
| Yes | 407 (20.4%) | 8 (8.9%) | |
| **Weight Loss** | | | |
| No | 1457 (73.0%) | 46 (51.1%) | <0.001 |
| Yes | 540 (27.0%) | 44 (48.9%) | |
| **Urinary catheter at baseline** | | | |
| Yes | 1162 (58.2%) | 62 (68.9%) | 0.043 |
| No | 835 (41.8%) | 28 (31.1%) | |
| **Feeding tube at baseline** | | | |
| Yes | 73 (3.7%) | 7 (7.8%) | 0.046 |
| No | 1924 (96.3%) | 83 (92.2%) | |
| **Ventilator at baseline** | | | |
| Yes | 760 (38.1%) | 42 (46.7%) | 0.1 |
| No | 1237 (61.9%) | 48 (53.3%) | |
| **Central venous catheter at baseline** | | | |
| Yes | 878 (44.0%) | 37 (41.1%) | 0.593 |
| No | 1119 (56.0%) | 53 (58.9%) | |

Antibiotics

| | Colonized (N=1997) | Case (N=90) | ***P*** |
|---|---|---|---|
| **Prior use of clindamycin** | | | |
| 0 | 1969 (98.6%) | 87 (96.7%) | 0.138 |
| 1 | 28 (1.4%) | 3 (3.3%) | |
| **Prior use of cephalosporins** | | | |
| No | 1806 (90.4%) | 71 (78.9%) | <0.001 |
| Yes | 191 (9.6%) | 19 (21.1%) | |
| **Prior use of penicillins** | | | |
| No | 1756 (87.9%) | 62 (68.9%) | <0.001 |
| Yes | 241 (12.1%) | 28 (31.1%) | |
| **Prior use of quinolones** | | | |
| 0 | 1966 (98.4%) | 85 (94.4%) | 0.004 |
| 1 | 31 (1.6%) | 5 (5.6%) | |
| **Prior use of carbapenems** | | | |
| 0 | 1978 (99.0%) | 80 (88.9%) | <0.001 |
| 1 | 19 (1.0%) | 10 (11.1%) | |
| **Prior use of monobactams** | | | |
| 0 | 1988 (99.5%) | 87 (96.7%) | <0.001 |
| 1 | 9 (0.5%) | 3 (3.3%) | |
| **Prior use of aminoglycosides** | | | |
| No | 1951 (97.7%) | 74 (82.2%) | <0.001 |
| Yes | 46 (2.3%) | 16 (17.8%) | |
| **Prior use of macrolides** | | | |
| 0 | 1942 (97.2%) | 82 (91.1%) | <0.001 |
| 1 | 55 (2.8%) | 8 (8.9%) | |
| **Prior use of tetracyclines** | | | |
| 0 | 1985 (99.4%) | 88 (97.8%) | 0.065 |
| 1 | 12 (0.6%) | 2 (2.2%) | |
| **Prior use of daptomycin** | | | |
| 0 | 1992 (99.7%) | 90 (100%) | 0.635 |

| | **Colonized (N=1997)** | **Case (N=90)** | ***P*** |
|---|---|---|---|
| 1 | 5 (0.3%) | 0 (0%) | |
| **Prior use of rifamycins** | | | |
| 0 | 1970 (98.6%) | 87 (96.7%) | 0.122 |
| 1 | 27 (1.4%) | 3 (3.3%) | |
| **Prior use of polymyxin** | | | |
| 0 | 1994 (99.8%) | 90 (100%) | 0.713 |
| 1 | 3 (0.2%) | 0 (0%) | |
| **Prior use of fosfomycin** | | | |
| 0 | 1976 (98.9%) | 89 (98.9%) | 0.957 |
| 1 | 21 (1.1%) | 1 (1.1%) | |
| **Prior use of nitrofurantoin** | | | |
| 0 | 1990 (99.6%) | 89 (98.9%) | 0.253 |
| 1 | 7 (0.4%) | 1 (1.1%) | |
| **Prior use of antituberculosis medications** | | | |
| 0 | 1992 (99.7%) | 89 (98.9%) | 0.136 |
| 1 | 5 (0.3%) | 1 (1.1%) | |

Other Medications

| | Colonized (N=1997) | Case (N=90) | *P* |
|---|---|---|---|
| **Prior use of immunosuppressive medications** | | | |
| 0 | 1926 (96.4%) | 84 (93.3%) | 0.126 |
| 1 | 71 (3.6%) | 6 (6.7%) | |
| **Prior use of diuretics** | | | |
| No | 1684 (84.3%) | 58 (64.4%) | <0.001 |
| Yes | 313 (15.7%) | 32 (35.6%) | |
| **Prior use of hypoglycemics** | | | |
| 0 | 1975 (98.9%) | 90 (100%) | 0.317 |
| 1 | 22 (1.1%) | 0 (0%) | |
| **Prior use of proton pump inhibitors** | | | |
| No | 1637 (82.0%) | 58 (64.4%) | <0.001 |
| Yes | 360 (18.0%) | 32 (35.6%) | |
| **Prior use of immunoglobulin** | | | |
| 0 | 1990 (99.6%) | 88 (97.8%) | 0.008 |
| 1 | 7 (0.4%) | 2 (2.2%) | |
| **Hemodialysis** | | | |
| 0 | 1993 (99.8%) | 89 (98.9%) | 0.084 |
| 1 | 4 (0.2%) | 1 (1.1%) | |
| **Nicotine use** | | | |
| 0 | 1972 (98.7%) | 88 (97.8%) | 0.426 |
| 1 | 25 (1.3%) | 2 (2.2%) | |
| **Prior use of vitamin D** | | | |
| No | 1832 (91.7%) | 69 (76.7%) | <0.001 |
| Yes | 165 (8.3%) | 21 (23.3%) | |
| **Prior use of angiotensin blockers** | | | |
| 0 | 1955 (97.9%) | 89 (98.9%) | 0.517 |
| 1 | 42 (2.1%) | 1 (1.1%) | |
| **Prior use of pressors/inotropes** | | | |
| No | 1854 (92.8%) | 66 (73.3%) | <0.001 |

| | Colonized (N=1997) | Case (N=90) | *P* |
|---|---|---|---|
| Yes | 143 (7.2%) | 24 (26.7%) | |
| **Prior use of antidepressants/antipsychotics** | | | |
| No | 1738 (87.0%) | 66 (73.3%) | <0.001 |
| Yes | 259 (13.0%) | 24 (26.7%) | |
| **Prior use of histamine antagonists** | | | |
| No | 1753 (87.8%) | 68 (75.6%) | <0.001 |
| Yes | 244 (12.2%) | 22 (24.4%) | |
| **Prior high risk antibiotic use** | | | |
| No | 1699 (85.1%) | 56 (62.2%) | <0.001 |
| Yes | 298 (14.9%) | 34 (37.8%) | |

Laboratory Results

| | Colonized (N=1997) | Case (N=90) | *P* |
|---|---|---|---|
| **Baseline circulating WBC (thousands of cells/microliter)** | | | |
| Mean (SD) | 6.48 (5.64) | 5.60 (3.78) | 0.037 |
| Median [Min, Max] | 6.00 [0.0900, 160] | 5.50 [0.0900, 15.5] | |
| Missing | 14 (0.7%) | 0 (0%) | |
| **Maximum circulating WBC (thousands of cells/microliter)** | | | |
| Mean (SD) | 16.6 (16.9) | 22.1 (18.9) | 0.008 |
| Median [Min, Max] | 13.6 [0.100, 322] | 18.9 [0.200, 143] | |
| Missing | 14 (0.7%) | 0 (0%) | |
| **Baseline serum hemoglobin (g/dL)** | | | |
| Mean (SD) | 8.74 (2.30) | 7.58 (1.79) | <0.001 |
| Median [Min, Max] | 8.30 [3.60, 17.4] | 7.00 [4.70, 13.4] | |
| Missing | 14 (0.7%) | 0 (0%) | |
| **Maximum serum hemoglobin (g/dL)** | | | |
| Mean (SD) | 11.7 (2.21) | 11.3 (1.97) | 0.067 |
| Median [Min, Max] | 11.4 [5.60, 19.7] | 10.7 [8.30, 17.9] | |
| Missing | 14 (0.7%) | 0 (0%) | |
| **Baseline circulating platelets (thousands of cells/microliter)** | | | |
| Mean (SD) | 145 (97.9) | 122 (94.0) | 0.023 |
| Median [Min, Max] | 134 [0, 627] | 107 [1.00, 380] | |
| Missing | 14 (0.7%) | 0 (0%) | |
| **Maximum circulating platelets (thousands of cells/microliter)** | | | |
| Mean (SD) | 283 (175) | 318 (184) | 0.082 |
| Median [Min, Max] | 249 [16.0, 2290] | 301 [27.0, 947] | |

| | Colonized (N=1997) | Case (N=90) | *P* |
|---|---|---|---|
| Missing | 14 (0.7%) | 0 (0%) | |
| **Baseline serum creatinine (mg/dL)** | | | |
| Mean (SD) | 0.870 (0.781) | 0.772 (0.495) | 0.078 |
| Median [Min, Max] | 0.700 [0.0900, 15.8] | 0.660 [0.160, 3.61] | |
| Missing | 19 (1.0%) | 0 (0%) | |
| **Maximum serum creatinine (mg/dL)** | | | |
| Mean (SD) | 1.67 (1.68) | 2.13 (1.67) | 0.013 |
| Median [Min, Max] | 1.08 [0.110, 19.1] | 1.54 [0.450, 8.39] | |
| Missing | 19 (1.0%) | 0 (0%) | |
| **Baseline serum albumin (g/dL)** | | | |
| Mean (SD) | 2.98 (0.750) | 2.58 (0.659) | <0.001 |
| Median [Min, Max] | 3.00 [0.0900, 5.20] | 2.50 [1.30, 4.10] | |
| Missing | 188 (9.4%) | 4 (4.4%) | |
| **Baseline serum albumin (categorical)** | | | |
| <2.5 g/dL | 353 (17.7%) | 39 (43.3%) | <0.001 |
| >=2.5 g/dL | 1456 (72.9%) | 47 (52.2%) | |
| Missing | 188 (9.4%) | 4 (4.4%) | |
| **Maximum serum albumin (g/dL)** | | | |
| Mean (SD) | 8.30 (98.2) | 3.55 (0.567) | 0.04 |
| Median [Min, Max] | 3.70 [1.50, 2540] | 3.60 [2.30, 4.90] | |
| Missing | 188 (9.4%) | 4 (4.4%) | |
| **Baseline serum protein (g/dL)** | | | |
| Mean (SD) | 5.22 (0.992) | 4.81 (0.987) | <0.001 |
| Median [Min, Max] | 5.10 [1.90, 9.10] | 4.60 [2.80, 7.90] | |
| Missing | 200 (10.0%) | 5 (5.6%) | |
| **Maximum serum protein (g/dL)** | | | |
| Mean (SD) | 6.28 (1.02) | 6.37 (1.04) | 0.441 |

| | Colonized (N=1997) | Case (N=90) | ***P*** |
|---|---|---|---|
| Median [Min, Max] | 6.20 [3.30, 11.2] | 6.30 [4.20, 9.50] | |
| Missing | 200 (10.0%) | 5 (5.6%) | |
| **Baseline serum glucose (mg/dL)** | | | |
| Mean (SD) | 89.7 (25.2) | 81.1 (25.9) | 0.003 |
| Median [Min, Max] | 87.0 [26.0, 278] | 79.5 [15.0, 164] | |
| Missing | 19 (1.0%) | 0 (0%) | |
| **Maximum serum gluconse (mg/dL)** | | | |
| Mean (SD) | 192 (91.3) | 227 (104) | 0.003 |
| Median [Min, Max] | 169 [65.0, 1170] | 193 [66.0, 596] | |
| Missing | 19 (1.0%) | 0 (0%) | |
| **Baseline hemoglobin A1C (%)** | | | |
| Mean (SD) | 6.49 (1.64) | 6.43 (1.54) | 0.892 |
| Median [Min, Max] | 6.00 [3.60, 13.7] | 6.00 [4.30, 9.40] | |
| Missing | 1721 (86.2%) | 76 (84.4%) | |
| **Maximum hemoglobin A1C (%)** | | | |
| Mean (SD) | 6.50 (1.64) | 6.43 (1.54) | 0.867 |
| Median [Min, Max] | 6.10 [3.60, 13.7] | 6.00 [4.30, 9.40] | |
| Missing | 1721 (86.2%) | 76 (84.4%) | |
